# New tests for trials of very few patients using longitudinal data - a case-study in Autosomal Recessive Cerebellar Ataxias

**DOI:** 10.64898/2026.08.28.26361588

**Authors:** Niels Hendrickx, France Mentré, Mats O. Karlsson, Andrew C. Hooker, Andreas Traschütz, Rebecca Schüle, PROSPAX Consortium, EVIDENCE-RND Consortium, Matthis Synofzik, Emmanuelle Comets

**Affiliations:** Université Paris Cité et Université Sorbonne Paris Nord, Inserm, IAME, F-75018 Paris, France; Department of Pharmacy, Pharmacometrics Research Group, Uppsala University, Uppsala, Sweden; Division Translational Genomics of Neurodegenerative Diseases, Hertie Institute for Clinical Brain Research (HIH), University of Tübingen, Tübingen, Germany; German Center for Neurodegenerative Diseases, (DZNE), Tübingen, Germany; Department of Neurology and Epileptology, Hertie Institute for Clinical Brain Research (HIH) and Center of Neurology, University of Tübingen, Tübingen, Germany; Department of Neurology, Division of Neurodegenerative Diseases and Movement Disorders, Heidelberg University Hospital and Faculty of Medicine, Heidelberg, Germany; Center for Neurology and Hertie Institute for Clinical Brain Research, University of Tübingen, Tübingen, Germany; Univ Rennes, Inserm, EHESP, Irset - UMR S 1085, 35000, Rennes, France

**Keywords:** N-of-1, N-of-few, NLMEM, longitudinal modelling, rare diseases, clinical trial design, disease progression

## Abstract

We propose two new tests to detect drug effects (DE) in trials of one to very few patients followed during two periods (before and after initiation of a treatment). Both methods use longitudinal natural history data to inform the estimation of each patient’s DE. The first method uses a non-linear mixed effect model (NLMEM) reflecting an expected natural history with a hypothetical drug effect, to estimate the Conditional Distribution of the Drug Effect (CDDE). The second method trains a Pareto Depth Analysis (PDA) algorithm, a machine learning–based approach based on outlier detection, that we implement using data simulated under the NLMEM. We evaluated the two tests with a simulation study. We used data from the PROSPAX study in Autosomal Recessive Cerebellar Ataxias (ARCAs, to derive a NLMEM for the Scale for the Assessment and Rating of Ataxia score. The CDDE method provided controlled type I error and, in some scenarios, adequate corrected power, though sensitivity analyses showed vulnerability to misspecification. The PDA method demonstrated lower statistical power except with high score precision. These results highlight different strategies for quantifying treatment effects in ultra-rare, patient-specific trials. They can inform methodological design for future ARCA precision therapies.

## 1 Introduction

Autosomal Recessive Cerebellar Ataxias (ARCAs) are ultra-rare, progressive neurodegenerative disorders that primarily affect the cerebellum but also cause multi-systemic involvement. Clinical manifestations include gait and balance disturbances, dysarthria, and impaired fine motor coordination [1]. More than 100 genotypes have been described [1], with onset typically occurring in childhood or early adulthood. In the absence of a universal biomarker, disease severity must be measured using clinical composite scales such as the Scale for the Assessment and Rating of Ataxia (SARA) [2]. The FDA has recommended leveraging natural history cohorts and patient registries to guide trial design, refine inclusion criteria, and define primary outcomes [3, 4]. Although no disease-modifying therapies are currently approved for most ARCAs, the monogenic nature of ARCAs makes them strong candidates for targeted molecular interventions, including gene therapy and antisense oligonucleotide (ASO) approaches [5]. Several such therapies aiming to slow disease progression are already under development. However, because gene therapy is inherently individualized, conventional randomized controlled trial (RCT) designs are not feasible. Instead, evaluation often relies on trials of very few patients [6]. In addition, conventional N-of-1 crossover designs are unsuitable in this context, as the therapies under investigation are intended to be disease-modifying rather than purely symptomatic.

In this work, we propose two tests for detecting disease-modifying treatment effects in trials of one or very few patients, integrating natural history data through longitudinal models. Both approaches assume that patients are first observed untreated, for instance as they are included in a registry, providing repeated measurements of an outcome of interest, before entering a treatment period with observations of the same outcome of interest. Both approaches also assume that a Non-Linear Mixed-Effect Model (NLMEM) was established on the natural history data, with the parameter distribution. The first test, called the Conditional Distribution of the Drug Effect (CDDE), is a model-based approach based on individual Bayesian regression, using a NLMEM of the natural history of the disease in the absence of treatment and on an assumption of the treatment effect (on one or several parameters) to derive conditional distributions for the individual parameters. The second test, called Pareto Depth Analysis (PDA), is an approach based on a machine learning algorithm for outlier detection [7], as well as to sort multivariate strategies in genetic algorithms [8]. Our approach is trained on data from untreated patients simulated from the NLMEM of the natural history data. Features of the progression are extracted from the longitudinal data such that patients in the trial would be considered outliers compared to the untreated patients if the DE has an effect.

We evaluated the two proposed tests on a simulation study with several scenarios, based on a NLMEM of the SARA score built on the PROSPAX study [9]. We considered N-of-1 and N-of-5 trials simulated with no Drug Effect (DE) to assess type I error and moderate or high DE to assess the corrected power. Finally, to evaluate robustness, we conducted a sensitivity analysis of the CDDE method with respect to the misspecification of the model.

We first present the two statistical tests, with the trial design and assumptions, then the simulation study that evaluates the performance of the two tests.

## 2 New tests for trials of very few patients

### 2.1 Trial design and assumption

We consider trials where a group of N patients (with N small, possibly N=1) are followed over two periods. Patients, included at time *t*_0,*i*_ (where *i* = 1…*N* denotes patient *i*) after disease onset, are first untreated for a duration *T*_1_ then treated for a duration *T*_2_. We assume that repeated observations *y*_*ij*_ (*j*=1…*n*_*i*_) of a continuous disease score are collected in each period. Our objective is to detect a treatment effect modifying the evolution of the disease in the second period compared to the first.

We also assume that the evolution of the disease on natural history is described using a NLMEM characterised by population parameters *θ*_*pop*_ where *θ*_*pop*_ = (*µ*, Ω, *σ*) denotes the population parameters in the absence of treatment.

Denoting *ψ*_*i*_′ the vector of parameters for subject *i*^′^ in the natural history study, we assume the following model for an observation 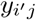 recorded in individual *i*^′^ at time 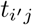 after disease onset with no treatment:

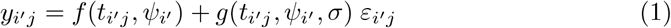

With 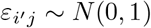, where *f* denotes the model function and *g* describes the variability of the residual error, depending on parameters *σ*. In the NLMEM, the individual parameters are modelled as:

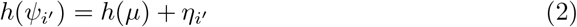

where *h* denotes a transformation of the individual parameters to a multivariate normal space for the random effects *η*_*i*_′, which are assumed to be drawn in *N* (0, Ω).

A graph representing an example of individual disease progression can be found in Figure 1.

**Fig. 1:**
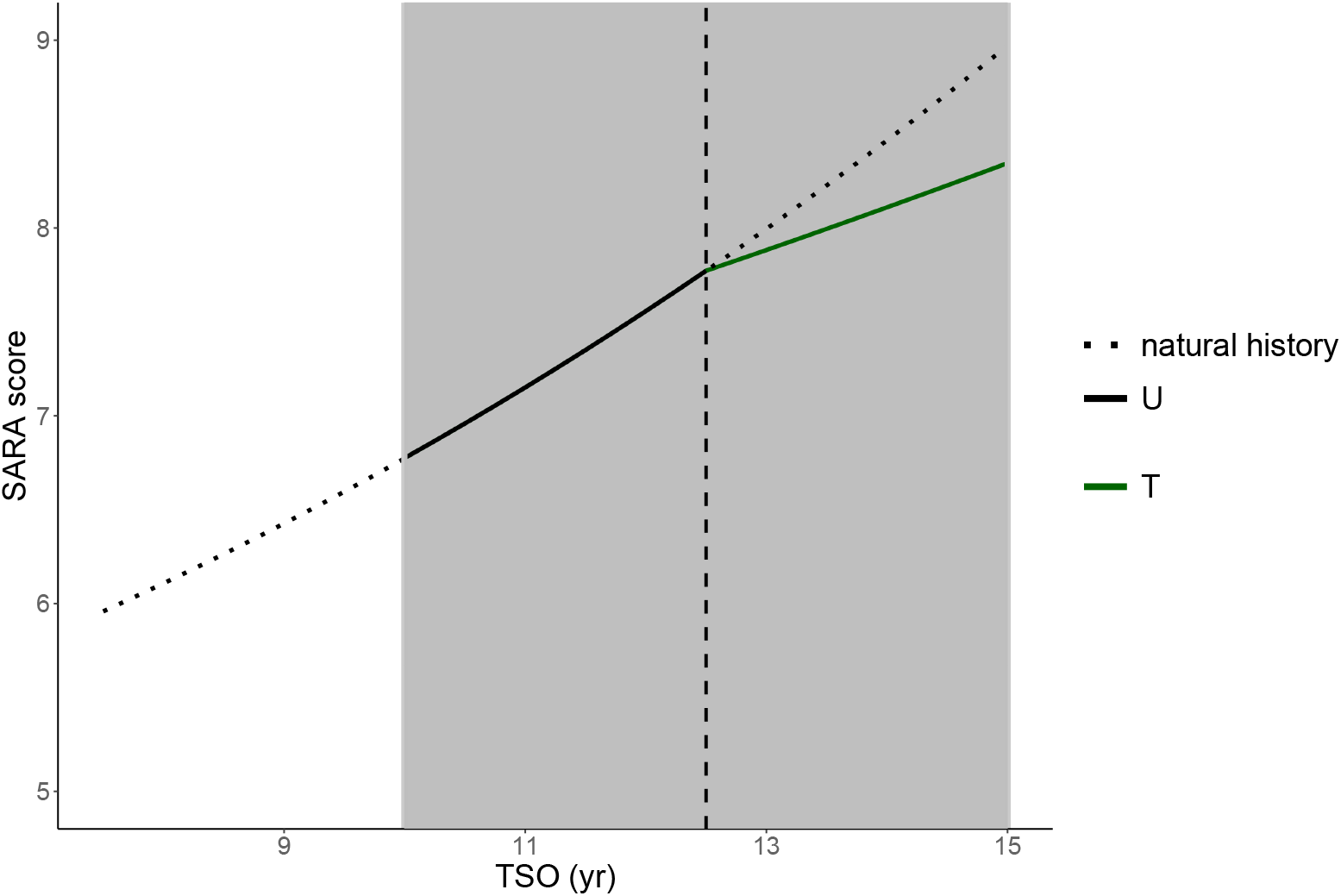
Simulated example of an individual trajectory during an N-of-1 trial, the shaded area corresponds to the trial period, the U (black solid line) corresponds to the untreated period and the T (green solid line) corresponds to the treated period, with the vertical dashed line delineating the beginning of the treatment period. The dotted line shows the trajectory before the observation period and the natural history during the second period if the patient remained untreated.

### 2.2 Conditional distribution of the drug effect (CDDE)

In the CDDE approach, the structure of the NLMEM developed on natural history data is complemented by adding a Drug Effect (DE) parameter, modelled using a normal distribution with a mean value *µ*_*DE*_ and a standard deviation *ω*_*DE*_. The full model is then characterised by *θ* = {*θ*_*pop*_, *θ*_*DE*_ }, with *θ*_*DE*_ = (*µ*_*DE*_, *ω*_*DE*_). In this model, the individual parameters are modelled as:

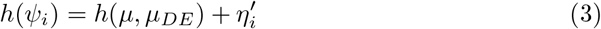

where the augmented random effects 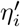 are now drawn in a multivariate normal distribution with variance-covariance matrix 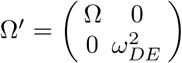.

To define our test, we use the population model and parameters as priors to estimate the conditional distribution of the individual parameters *ψ*_*i*_ for the i^*th*^ subject by an MCMC procedure coupled with an accept-reject algorithm [10], using the Metropolis-Hasting algorithm. During the untreated period, *θ*_*pop*_ can reflect a prior analysis of natural history data, using previously estimated values. For the drug effect, we assume a mean drug effect of 0 and we set a large variance 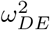. Denoting *ψ*_*DE,i*_ the DE component of *ψ*_*i*_, representing the individual drug effect in subject *i*, and *y* = (*y*_1_, …, *y*_*N*_ ), we test for the significance of the drug effect in the *N* subjects using the marginal conditional distribution *p*(*ψ*_*DE*_ *y*) where *ψ*_*DE*_ = {*ψ*_*DE,i*}*i*=1…*N*_ . We define the null hypothesis (*H*_0_) and alternative hypothesis (*H*_1_) as follows:

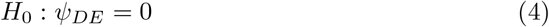

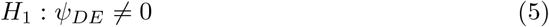

Under the assumption that the conditional distribution is normal, we can test the significance of *ψ*_*DE*_ using a multivariate Wald test with *N* degrees of freedom using the mean and variance of the conditional distribution of each *ψ*_*DE,i*_ (since we assume each patient is independent). Alternatively, we can also test the significance of *ψ*_*DE*_ by building a multivariate 95% Credibility Region using Highest Density Regions [11] (CR-HDR), which we develop in Appendix B.

### 2.3 Pareto depth analysis (PDA)

In this method, the NLMEM developed on natural history data is first used to simulate *P* untreated patients, included at times *t*_0,*p*_ (*p* = 1…*P* ) after disease onset, with the same design as the patients in the trial, to extract features summarising natural disease progression. We choose as features the observed slopes for both treatment periods for each patient, fitting the following piecewise linear model to an observation *y*_*pj*_ taken at time *t*_*pj*_ after inclusion in the study:

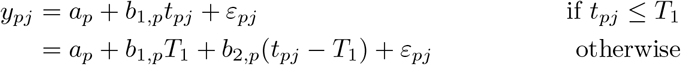

with *ε*_*pj*_ ∼ *N* (0, *σ*). We define the features as the 2-vectors *B*_*p*_ = (*b*_1,*p*_, *b*_2,*p*_). For an untreated patient, we expect that *b*_1,*p*_ = *b*_2,*p*_.

The dataset {*B*_*p*_} _*p*=1,…,*P*_ is split into two subsets, *B*_*train*_ (matrix of dimension *P*_*train*_ × 2) to train a machine-learning algorithm, and *B*_*test*_ (matrix of dimension *P*_*test*_ × 2), to calibrate it. Here, we propose to compute Pareto Fronts on *B*_*train*_ through a procedure proposed by [7] and described in detail in appendix C. Briefly, we first compute the element-wise distance between each vector of features in the training set, and use these to iteratively define sets of non-dominated elements. These constitute Pareto Fronts (ℱ_*l*=1…*L*_) representing the “typical” distances between the features in the training set. The depth *e*_*p*_ for an individual *p* with features *B*_*p*_ ∈ *B*_*train*_ is defined as the lowest front *l* not strictly dominating *B*_*p*_.

For an individual *p*^′^ in *B*_*test*_, we compute the mean depth as the mean of the depths of its *s*_*p*_′ nearest neighbours in *B*_*train*_.

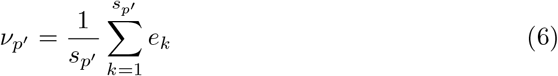

We then estimate a threshold *ρ* such that, for any *N* subjects 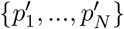 *B*_*test*_, we have 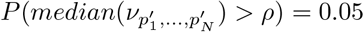.

Finally, we define a test for the N subjects in the trial by computing their mean depth as above, considering the drug effect to be significant if *median*(*ν*_*i*=1…*N*_ ) *> ρ*.

Figure 2 summarises the workflow of the PDA analysis.

**Fig. 2:**
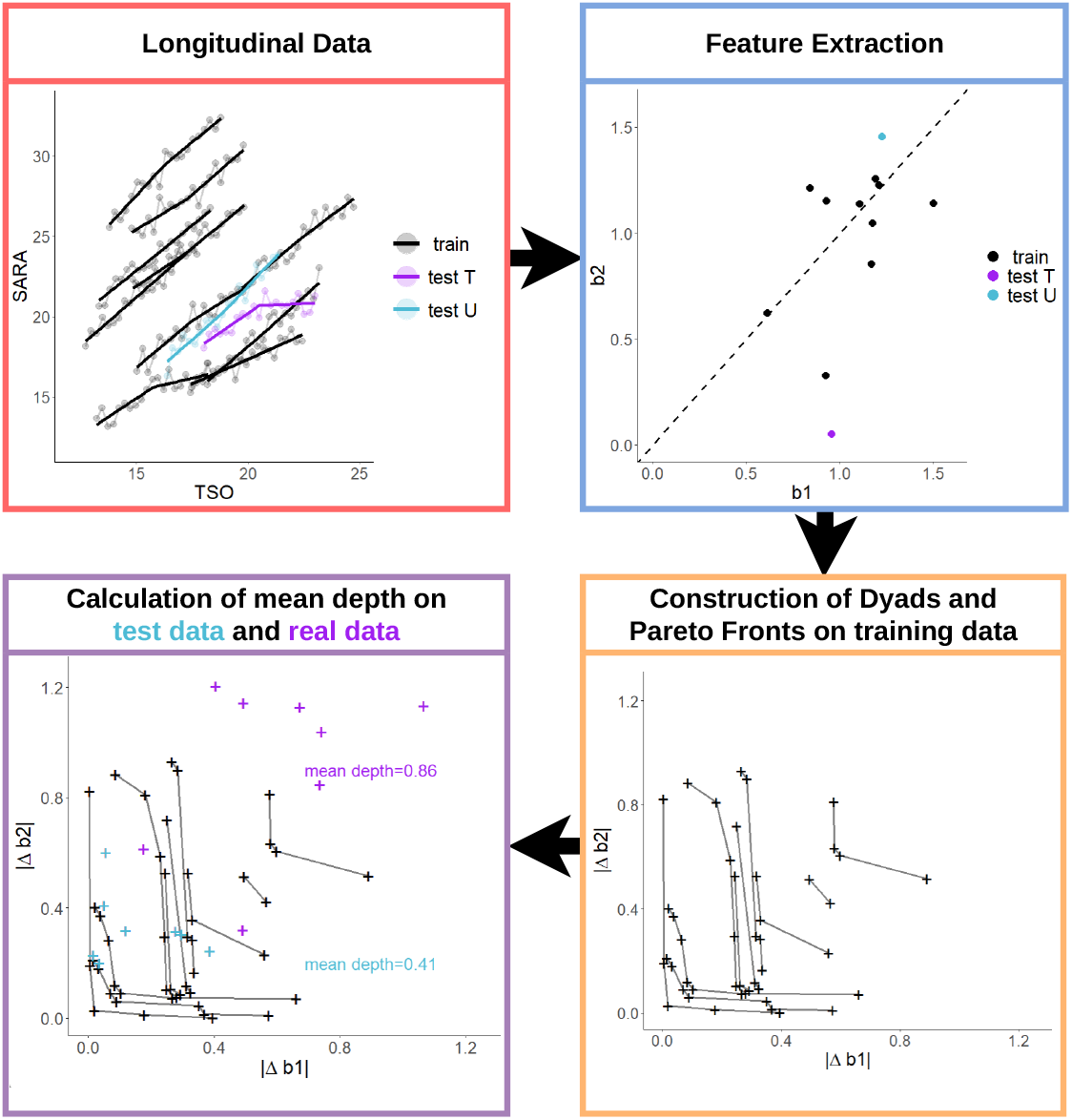
Flowchart representing the workflow of the PDA analysis, representing 10 patients in the training dataset (in black) simulated without DE, 1 patient in the calibration (in cyan) also simulated without DE and 1 test patient (in magenta), simulated with DE (details of the simulation models are given in section 3).

## 3 Evaluation by a simulation study

The two proposed tests were evaluated with a simulation study to quantify the type I error and power of the detection of a DE in trials of very few patients.

### 3.1 Simulation models

For this paper, we simulated data using a NLMEM developed to describe the evolution of total SARA scores in patients from the PROSPAX natural history study (NCT04297891) [9]. These patients, enrolled in Europe and in Canada, had genetically confirmed diagnosis of ARSACS or SPG7, two Autosomal Recessive Cerebellar Ataxias. The disease severity was measured using the SARA score [2], a composite score comprised of 8 clinical sub-items evaluating speech, gait, stance sitting and fine motor movement. The main recorded characteristics at inclusion were age, sex, Age Of Onset (AOO), SARA score. Only patients with at least one SARA score and non-missing age of onset were selected, yielding a total of 81 patients with the ARSACS genotype and 113 patients with the SPG7 genotypes, most of whom had at least two visits. Details of the dataset, model selection and estimation results are given in appendix D. The SARA score at time *t* after the disease onset, considered to be the time since disease onset (TSO), in the absence of treatment was modelled as a continuous variable using a 4-parameter logistic model previously developed a similar population using registry data [12]:

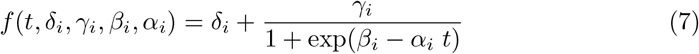

The parameters were considered to be log-normally distributed in the population and the residual error model was homoscedastic (*g* = *σ* in equation 1). The resulting NLMEM was fit using the *saemix* package [13] in R 4.1.2 [14] to the subset of patients with the ARSACS genotype, yielding estimated parameters reported in the first column of table 1. We then introduced a hypothetical DE on the *α* parameter, assuming it slowed disease progression during the second part of the trial, leading to the following simulation model for the two periods, with *t* the time since disease onset:

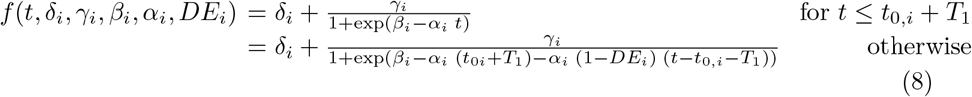

where *t*_0*i*_ denotes the time since disease onset at the beginning of the trial, and *DE*_*i*_ = 0 for untreated patients.

**Table 1:** Parameters and their Relative Standard Errors (RSE) for the different models used in the simulation and the analyses.

| Parameter | Logistic model |  |  |  | Linear model |  |  |
| --- | --- | --- | --- | --- | --- | --- | --- |
|  | ARSACS |  | SPG7 |  | ARSACS |  |  |
|  | Value | RSE (%) | Value | RSE (%) | Parameter | Value | RSE (%) |
| $\delta$ | 7.65 | 36 | 0* | - | $a$ | 16.41 | 4 |
| $\gamma$ | 27.38 | 27 | 37.99 | 17 | $\beta_{TSO_{bl},a}$ | 0.45 | 11 |
| $\beta$ | 3.0 | 34 | 1.55 | 11 | $b \text{ (yr}^{-1}\text{)}$ | 0.64 | 32 |
| $\alpha \text{ (yr}^{-1}\text{)}$ | 0.083 | 40 | 0.04 | 20 | $\beta_{TSO_{bl},b} \text{ (yr}^{-2}\text{)}$ | -0.031 | 59 |
| $\omega_\delta$ | 0.30 | 42 | 0* | - | $\omega_a$ | 5.0 | 9 |
| $\omega_\gamma$ | 0* | - | 0* | - | $\omega_b$ | 1.0 | 23 |
| $\omega_\beta$ | 0.26 | 28 | 0.25 | 14 | | | |
| $\omega_\alpha$ | 0.15 | 77 | 0.45 | 18 | | | |
| $\sigma$ | 1.84 | 7 | 1.56 | 5 | $\sigma$ | 1.50 | 9 |

\* fixed parameters

### 3.2 Simulation scenarios

We defined 12 simulation scenarios combining the following parameters: (i) disease progression rate (*α* = 0.08 as estimated in ARSACS patients or *α* = 0.16 for fast progression); (ii) residual error (*σ* = 1.84 as estimated in ARSACS patients, or a lower value of 0.5 representing a more precise score); (iii) drug effect, with DE = 0 to assess type I error (H_0_), 0.5 or 1 to assess power (*H*_1_). The other parameters in the model were taken from the first column of table 1.

We performed two simulations for each scenario, *N* = 1 (N-of-1) and *N* = 5 (N-of-5). For each simulation, we generated 1000 replicates assuming *T*_1_ = *T*_2_ = 2.5 years and 4 visits per year (months 0, 3, 6 and 9). TSO at baseline (*t*_0,*i*_) were simulated uniformly between 20 − 30 years (for *α* = 0.08) or 10 − 20 years (for *α* = 0.16).

### 3.3 Implementation and evaluation

#### Implementation

The CDDE method was implemented in NONMEM 7.5.1 [15], using 1000 samples for the conditional distribution. The confidence regions for the CR-HDR test were obtained using all the samples for each patient. A first analysis was performed using for *θ*_*pop*_ the values used in the corresponding scenario and *θ*_*DE*_ = (0, 0.5, 1), and, for the estimation, the value *ω*^2^ = 25 was used as a large variance.

For the PDA method, in each scenario, 500 replicates simulated under *H*_0_ (*DE* = 0) were used to train the Pareto Front algorithm and the remaining 500 were used for calibration. The PDA approach was then applied to 500 replicates in each of the simulations under *H*_1_ (*DE* = 0.5 and *DE* = 1) to evaluate the power. The PDA method was implemented in Python 3.11.4.

#### Sensitivity analysis

We performed two sensitivity analyses on the simulations with *N* = 1 to evaluate the impact of model misspecification on the performance of the CDDE method. In the first, we consider the impact of prior parameter misspecification by using for *θ*_*pop*_ the parameters in column 2 of table 1, which were obtained by analysing the SPG7 subset with the same procedure as described above (see appendix D). In the second, we considered the impact of structural model misspecification, by using a linear mixed effect model (LMEM) instead of the logistic model. The structural model assumed that treatment effect slowed disease progression by changing the slope:

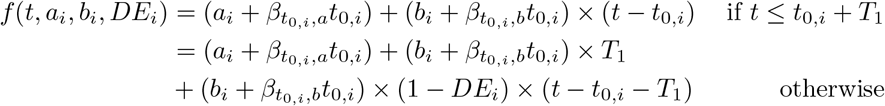

where *a*_*i*_ and *b*_*i*_ represent the intercept and slope respectively. The corresponding *θ*_*pop*_ were obtained by fitting the LMEM to the subset of patients with the ARSACS genotype, assuming a log-normal distribution for *a*_*i*_, a normal distribution for *b*_*i*_, and are reported in the last column of table 1.

#### Evaluation

For both CDDE tests, the empirical type I error was calculated on each simulation scenario under *H*_0_, and used to correct the power for the simulations under *H*_1_ (choosing the significance threshold so that the type I error was 5%). For the PDA method, the type I error was 5% by calibration, and we computed the power in the simulations under *H*_1_.

### 3.4 Results

#### 3.4.1 CDDE method

The top two rows in Figure 3 show the type I error and the corrected power for all scenarios of the CDDE method using the CR-HDR and Wald tests, for N-of-1 and N-of-5 trials. The tables with the results can be found the supplementary tables S-2 and S-3. Both tests were comparable for all scenarios (difference of a few percent). Type I error was well controlled for all scenarios. For the N-of-1 trials, the corrected power was low for *DE* = 0.5 in all scenarios (*<* 60%). For *DE* = 1, we see that the corrected power remained low for a high *σ* (*<* 50%) but increased to more than 65% for a low *σ*. For the N-of-5 trials, there was an increase in power in all scenarios compared to the N-of-1 setting. Notably, for *DE* = 0.5 and low *σ*, the power was relatively high (*>* 60%). For *DE* = 1, the corrected power was higher than 85% with N-of-5 trials assuming fast progressing ataxias.

**Fig. 3:**
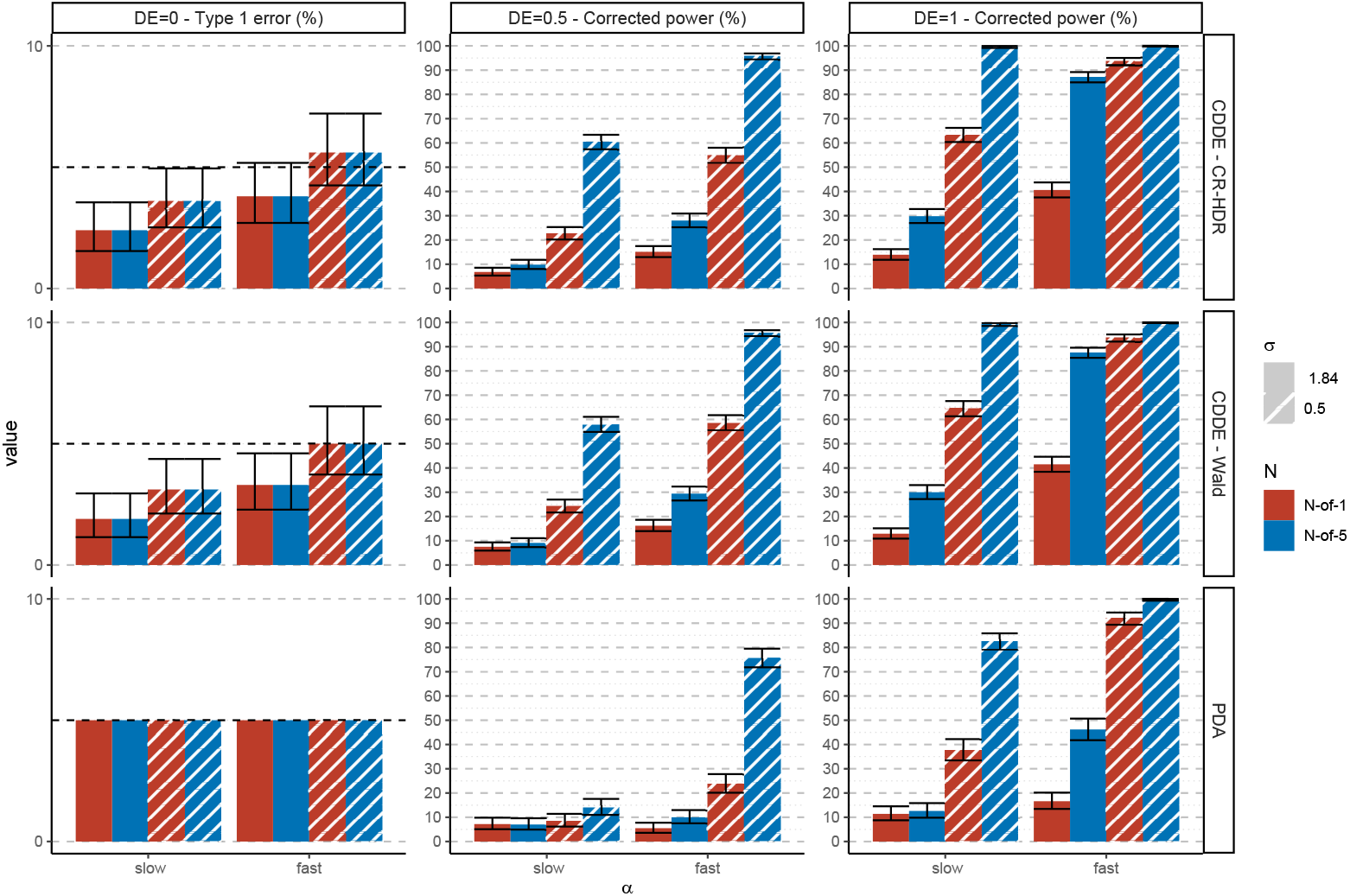
Type I error and power for slow and fast progression (*α*) of the CDDE, with the CR-HDR (top) and Wald (middle) tests, and PDA methods (bottom). The colour represents the number of patients (red=1, blue=5), solid bars represent the results for *σ* = 1.84 and dashed bars represent the results for *σ* = 0.5. Error bars correspond to the 95% confidence intervals of the powers and type I errors (calculated with 1000 replicates for the CDDE method and 500 replicates for the PDA method)

#### 3.4.2 Pareto Depth Analysis method

The last row in Figure 3 shows the type I error and corrected power for all scenarios for the PDA approach. By design, the type I error of this method is 5%, as the significance threshold is chosen by simulations under *H*_0_. For *DE* = 0.5, the power was very low in all scenarios (*<* 25% except the most advantageous scenario: N-of-5/fast progression/*σ*=0.5). For *DE* = 1, the power was still low for *σ* = 1.84 (*<* 50%, even with N-of-5). However, if *σ* = 0.5 and *DE* = 1, the corrected power reached 90% for N-of-1 and more than 99% corrected power in N-of-5 for fast progression. Compared to the CDDE method, the PDA method had lower power in all scenarios, especially with high magnitude of residual error. However, with small residual errors, the PDA method was almost as powerful as the CDDE method.

#### 3.4.3 Sensitivity analysis for CDDE

Figure 4 shows the type I error and corrected power in the sensitivity analysis for the CDDE method, investigating the impact of structural model misspecification (data simulated using the logistic and analysed with a linear model) and prior parameter misspecification (data simulated using parameters of the ARSACS genotype and analysed using parameters estimated on the SPG7 genotype). The tables with the results can be found the supplementary tables S-4 and S-5. For the prior parameter misspecification, the type 1 error was controlled with slow progression but was inflated for the fast progression scenarios. For the structural model misspecification, the type I error was controlled. We also see that the type I error inflation was greater for the CR-HDR test than the Wald test. For the slow progression scenarios, the corrected power of the prior parameter misspecification model was almost as high as the corrected power of the true (well specified) logistic model. For the fast progression scenarios, the structural misspecification model was more powerful than the prior parameter misspecification model in almost all scenarios.

**Fig. 4:**
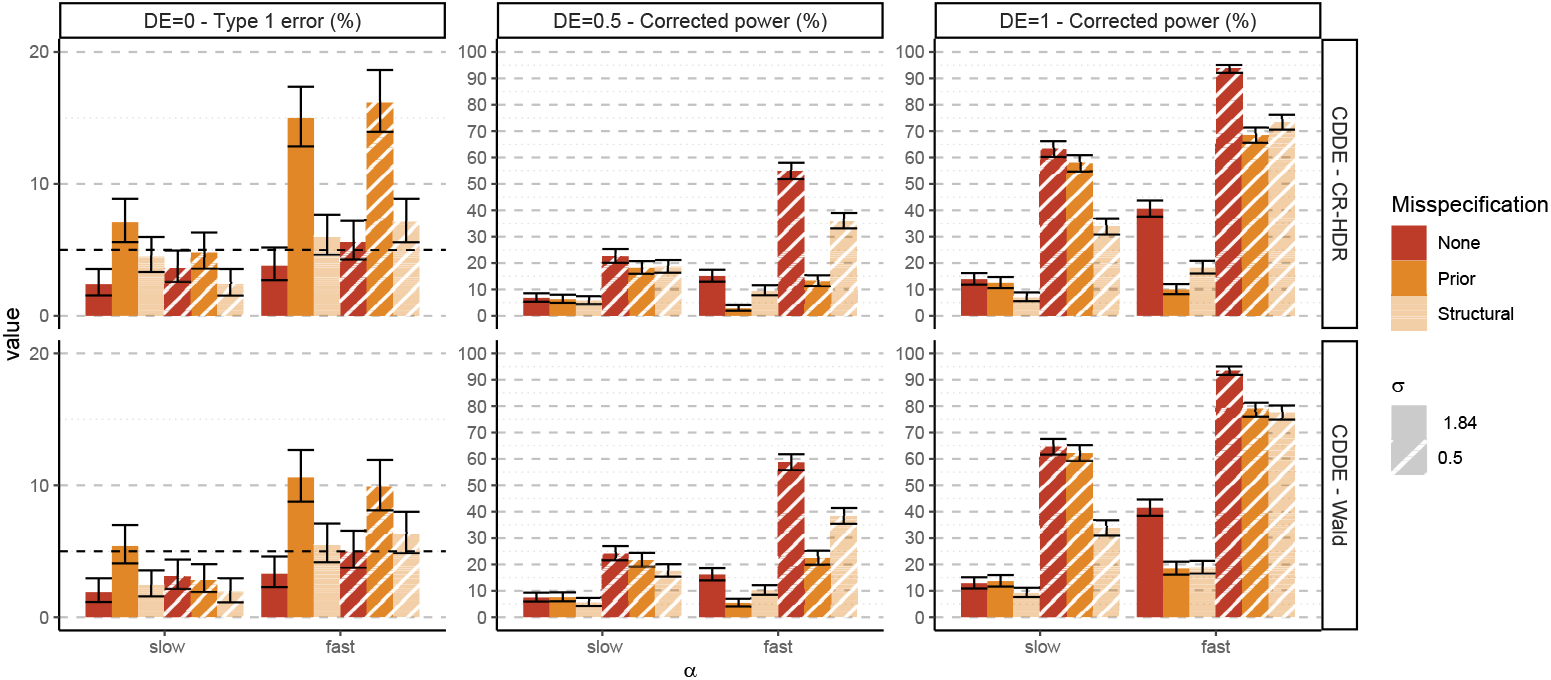
Type I error and corrected power for slow and fast progression (*α*) of the sensitivity analysis of the CDDE method (with the CR-HDR and Wald tests), with no misspecification (red), prior parameter misspecification (darker orange = priors of the logistic model estimated on the SPG7 genotype), or structural misspecification (lighter orange = data analysed with LMEM). Solid bars represent the results for *σ* = 1.84 and dashed bars represent the results for *σ* = 0.5. Error bars correspond to the 95% confidence intervals of the powers and type I errors (calculated over 1000 replicates).

## 4 Discussion

In this study, we developed and evaluated two approaches for detecting disease-modifying treatment effects in ultra-rare neurodegenerative diseases within trials of very few patients. The first is a model-based strategy using longitudinal regression (CDDE method), which incorporates parameters estimated from the natural history of disease progression data as priors for individual trajectories. It assumes a Drug Effect on one of the parameters of the model, with mean 0 and large variance, and the method then samples from the conditional distribution of the Drug Effect parameter for each patient in the trial. The second is the Pareto Depth Analysis method, a machine learning approach, that we implemented by training it on data simulated with a NLMEM to assess whether a treated patient deviates from external controls or not. Both proposed methods were built around N-of-1 trials, but they readily extend to N-of-few trials as an aggregate of N-of-1 trials. They were evaluated in N-of-1 and N-of-5 contexts using simulation studies, with a substantial increase in corrected power when increasing the number of subjects. We further examined the robustness of the CDDE method to structural misspecification (simulation with a logistic model analysed under a linear model) and to prior misspecification (simulation based on ARSACS parameters analysed with SPG7 parameters).

In the literature, Bayesian inference has been proposed to detect a drug effect in N-of-1 trials [16]. In that paper, the authors propose to assess DE using its posterior distribution, like in our CDDE approach, checking if 0 lies within its credible interval, like with our CR-HDR approach. However, they don’t explicitly define a method to calibrate the prior values, whereas, in this work, we propose an model-based method to leverage natural history data and define a global test for N-of-few trials. Hawksworth et al. [17] conducted a methodological review of the analysis of N-of-1 trials. This review highlights several points. Classical N-of-1 trials consist of a multiple crossover design, where patients can be randomised to a sequence of treatment. More than 65% of trials pooled several N-of-1 trials for the analysis, but the difference with a classical crossover trial is that the estimation of the DE is individual (whereas, in a classical crossover trial, an average DE is estimates). However, this review did not consider designs incorporating natural history data or information. N-of-1 trials were also used in rare and chronic diseases, but the direct application of such designs would be challenging in ARCAs for disease-modifying treatments (as the carry-over effect would not be negligible). Finally, statistical analysis methods showed a large heterogeneity, with Bayesian models, non-parametric tests, t-tests and regression models accounting for the methods used in over 70% of the studies.

In our simulation study, for the N-of-1 trials, it was possible to have more than 70% power using the CDDE method for the N-of-1 analysis. It however required favourable conditions, with a high signal to noise ratio, since evaluating disease-modifying treatments requires to observe the disease progression with sufficient accuracy to detect a change. We also assessed the sensitivity of the CDDE method to misspecification. Using priors obtained by estimating the model on another genotype (SPG7), type I error was conserved and corrected power was almost as high as for the well specified logistic model for the N-of-1 scenarios with slow progression, but the type 1 was inflated for fast progression. It may have helped that the corrected power for those scenarios was small to begin with, but it could also be due to the fact that, even if the fixed effects were estimated to different values for all 4 parameters, the standard deviation of the random effects were estimated to be larger in SPG7 compared to ARSACS (for *ω*_*α*_, it was estimated as 0.45 in SPG7 and 0.15 in ARSACS), so the prior would be less informative. In the fast progression case, the type I error was inflated for the prior parameter misspecification, most likely due to the fact that there were two layers of model misspecification. Put together, this reflects that the logistic model estimated on natural history data is generic and that it can handle mild model mis-specification. Another form of sensitivity analysis could be considered. The observed data from PROSPAX could be used as the untreated period of the N-of-1 trial, coupled with simulated trajectories for the trial period under treatment. We notice that both of our tests, the Wald test assuming the normality of the posterior distribution and the CR-HDR test, yielded the same power and type I error in all scenarios. It could be explained by the fact that, in our simulation settings, the model was well specified, the priors were calibrated around the true parameter values and that our sampling was quite rich (21 observations per patient), so we had a good estimation of the posterior distribution of the individual drug effect. With sparser sampling, we could expect the CR-HDR test to outperform the Wald test. In the prior misspecification scenario, the Wald test was more conservative than the CR-HDR test. Finally, the priors, which were estimated on the PROSPAX study, were very informative on the ARSACS genotype. In the context of a real study, one could inflate the variance of the priors on some parameters (especially *α*) to account for the uncertainty related to parameter estimation on the natural history data.

The Pareto Depth Analysis seems promising as a more data-driven approach to detect a drug effect in N-of-1 trials. As expected, this method was less powerful than the CDDE method in almost all scenarios, as the Pareto Depth analysis doesn’t make as many assumptions on the disease progression. In our implementation, simulated data was used to train and calibrate the model, making the method less data-driven and more model-based, but the drug effect agnostic approach of the feature selection and training makes it less sensitive to model misspecification as it doesn’t make assumptions on the structural model. A fully data-driven implementation would require large amounts of data allowing feature extraction (here, sufficiently rich samples during an duration similar to that of the trial to be able to extract the slopes in the linear model), which would be infeasible in rare diseases. For fast progression and low *σ*, PDA was almost as powerful as the CDDE method. In this work, the metric used to define what an outlier is was the slope during each treatment period. It was chosen as such because it was the easiest to implement, but the drawback is that we had to train the algorithm on simulated data that had the same design as treated patients, here observed for 5 years, with enough observations per period to identify both slopes. Other metrics were considered and evaluated. For example, one approach was to estimate all the individual parameters of the logistic model or a linear model and to build the distribution of individual parameters: *B*_*p*_ = *η*_*p*_. It has the advantage of not making any assumption on the drug effect or the design, meaning that the algorithm could be trained on observed, non treated patients (e.g. PROSPAX study), and tested on treated patients. There were, however, two challenges. First, assuming that DE is on *α*, since the *ω*_*α*_ is quite low (0.15), to accurately guess a DE of 0.5 without specifying DE in the model equation, the model would need to guess a random effect on *α* at more than 4 times the standard deviation of its prior, which is close to impossible. It means that the influence of DE is spread over all 4 parameters, making it more challenging to detect.

In a potential future study, the features could be enriched by adding covariates that could explain the variability in disease progression. As such, the biggest bottleneck for the PDA method would be finding a metric, based on the data, ideally independent of the design of the data or a model, and able to discriminate between untreated and treated patients. However, there are still challenges to implement it in practice. Even if features of disease progression were extracted from observed natural history data, the PDA method has no inherent methods to handle missing or incomplete data. Also this method requires a large amount of training data, which is not always available for rarer ARCA genotypes. In that case, one solution could be to pool and analyse several ARCA genotypes together, but that would also increase the variability and potentially dampen the signal. Finally, for both the CDDE and PDA methods, we did not investigate the impact of the design of the study (number of observation per patients, duration of each period, duration of the trial) in the simulation study. In fact, we considered an optimistic design as a proof of concept for the two proposed tests. The sensitivity of the power and type I error to the design characteristics could be investigated in a future study. In the context of a real study, power correction by the type 1 error inflation or deflation is not feasible in practice, which could impact the results.

Other approaches, or variants of the two proposed methods could have been used. First, in the CDDE method, a continuous model of the SARA score was chosen as in [12, 18], but other models could have been considered. Although, on the real data, linear models perform slightly worse than the logistic model, they are more robust to model misspecification as the disease progression is slow, so it would be a reasonable assumption. Alternatively, we could have considered a discrete model of the SARA score, such as an Item Response Theory model (IRT), which have shown good performance in ARCAs [19, 20]. Such models would allow us to model the SARA score with more granularity as each sub-score is modelled. Then, other, more standard statistical approaches could have been tested. For example, for the design we proposed, we tried fitting a piecewise linear (non mixed) model (one slope for each treatment period) on each patient and tested with a Wald test whether the 2 slopes were significantly different or not. The results were as expected in the sense that the type I error was controlled but the corrected power was lower than the CDDE in all scenarios, which makes sense because fewer assumptions are made with that approach. An extension of this work could be to use the Multivariate Exact Discrepancy (MED) metric [21]. MED is a metric of how adequate a model is to describe the evolution of a patient. The idea would be that, under the model estimated on the natural history, a treated patient would be misspecified and therefore they would be an outlier with regards to that metric. The upside of this metric is that no assumption is made on the drug effect, but the downside is that this metric wouldn’t differentiate between a “well-specified” treated patient and a “misspecified” untreated patient. Finally, one could design N-of-few trials as trials with external control, leveraging natural history data. For example, one could imagine enrolling 5-10 patients treated with a rich sampling design similar to our study and matching them (according to covariates at inclusion) with 50 untreated patients from natural history data using propensity score matching [22–24] to build an external control arm, to analyse the data as a randomised trial. In this work, with both approaches, we assumed that a patient in a trial, while under placebo, progresses in the same way as a patient in a natural history study, thereby neglecting the placebo effect. This assumption is debatable, especially since the primary endpoint is the SARA score, which measures symptom severity. This could imply an overestimation of the treatment efficacy. In a real clinical study, the placebo effect could depend on the experimental protocol and the nature of the intervention being tested. However, in Choi et al. [25], the authors performed a meta-analysis to quantify the placebo effect on the SARA score, in the context of randomised trials for Cerebellar Ataxias. Their conclusion was that, while there are trials where patients can be partial responders in the placebo group, there was no statistically significant placebo effect.

Ultra-rare diseases in general and ARCAs in particular pose specific difficulties for the conduct of clinical trials [12]. The patient population is both small and genetically heterogeneous, and natural history studies have shown that progression is generally slow and accompanied by substantial variability in SARA trajectories [18]. In a previous work, we developed a non-linear mixed effect model on ARCA patients, based on registry data [18], and used it to evaluate the power and type I error of randomised controlled trials of 40 to 100 patients in total [12]. However, for some ARCA genotypes, even the smaller sample sizes would be unfeasible, making it all the more relevant to develop tests for trials of very few patients. In fact, such designs are being considered in ARCAs and in ultra-rare neurodegenerative diseases in general [5]. In monogenic diseases, N-of-1 trials are suitable to evaluate the efficacy of individualised therapies such as gene therapies or antisense oligonucleotide therapies. The CDDE method we proposed would be compatible with the development process of such therapies. In the N-of-1 development roadmap [6], the authors explain that, to develop a N-of-1 treatment, the disease mechanism should first be understood, and then natural history data should be gathered. In our design, this could translate as the untreated period in the N-of-1 trial, which could help build a natural progression model of the disease. Then, the patient would be given the treatment and be monitored, which would be the second period of our N-of-1 trial. The long duration of the trial could be mitigated by including patients who were already monitored in a patient registry (for the initial 2.5 years without treatment). This would considerably reduce the duration of the trial but the impact of the number of observation per period and the duration of each period would need to be investigated. One could imagine a design where a patient would be monitored in a registry, without treatment, with an observation each year (as in PROSPAX), and then they would be monitored under treatment with a richer sampling schedule (4 observations per year for example). However, this work also quantified that, in order to have a good power with trials of one to five patients, fast disease progression rate, low residual error and long follow-up were required which could be a challenge for ARSACS patients. The question remains however at what point we could assume the asymptotic assumption is met and when we could use randomised trial approaches. In a previous study [12], we compared randomized trial designs of 100 and 40 patients, which had good performance. However, with trials of 20 patients, the type I error was inflated for all scenarios, so we could assume that our N-of-few approaches could also be applied at N=20.

## 5 Conclusion

In this work, we proposed two new methods to evaluate a disease-modifying treatment effect in trials of very few patients (down to one) for ultra-rare neurodegenerative diseases. The proposed methods leveraged natural history data in two ways: either by building a natural progression model to predict an individual drug effect, or by simulating observations of patients who were not treated as a reference to detect outliers in disease progression. Our analysis showed that, given suitable settings, N-of-1 trials, but preferably N-of-5 trials could be feasible with the proposed methods.

## Acknowledgements

This work was supported by members of the Evidence-RND consortium, which includes Alzahra Hamdan, Xiaomei Chen, Nicole Maria Heussen, Ralf-Dieter Hilgers, Thomas Klockgether, Yevgen Ryeznik, Oleksandr Sverdlov. This work was funded by the European Joint Programme on Rare Diseases (EJP RD) Joint Transnational Call 2019 for the EJP RD WP20 Innovation Statistics consortium “EVIDENCE-RND” focusing on “Innovative Statistical Methodologies to Improve Rare Diseases Clinical Trials in Limited Populations” under the EJP RD Grant Agreement (n°825575) (to M.K, R.S. and M.S.); as well as by the European Union, project European Rare Disease Research Alliance (ERDERA), GA n°101156595, funded under call HORIZON-HLTH-2023-DISEASE-07 (to M.S. R.S, and F.M.). Moreover, work in this project was supported by the Clinician Scientist programme “PRECISE.net” funded by the Else Kröner-Fresenius-Stiftung (to M.S., R.S. and A.T) and the Bundesministerium für Bildung und Forschung (BMBF) through funding for the TreatHSP network (grant 01GM2209A to R.S.), and by the European Health and Digital Executive Agency (HADEA) through funding for the European Rare Disease Research Alliance (ERDERA) (grant agreement 101156595), and the European Union via funding for the MSCA Doctoral Network Medicine Made to Measure (grant agreement #101120256 to RS), and RS is a member of the European Reference Network for Rare Neurological Diseases – Project ID 739510.

## Data availability statement

Data supporting the study findings are available on request.

## Ethical approval and informed consent statements

This study was conducted using data acquired within the multicenter project “An integrated multimodal progression chart in spastic ataxias” (PROSPAX) (ClinicalTrials.gov No. NCT04297891), approved by the local ethics committee of each center in accordance with the ethical standards of the institutional research committee and with the 1964 Helsinki Declaration and its later amendments. Written informed consent was obtained from each patient prior to enrollment.

## A PROSPAX Consortium author list

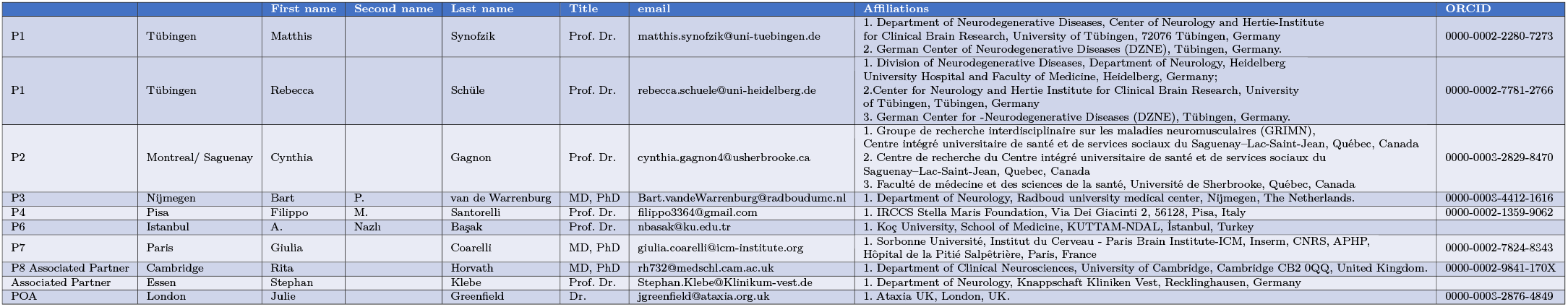

## B Credibility Region - Highest Density Region

In this section, we detail how to build a test based on credibility regions with highest density regions, assuming we are testing *N* patients.

We consider the vector of individual drug effects *ψ*_*DE*_, and we assume that for each patient *i* = 1…*N*, we have *S* (*s* = 1…*S*) Monte Carlo samples of the individual drug effect 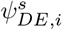.

Let *p*(·|*y*) denote the joint posterior density of *ψ*_*DE*_ and *p*_*i*_( |*y*_*i*_) the marginal posterior density of *ψ*_*DE,i*_ for individual *i*. We want to define a Confidence Region with risk *α, C*_1*−α*_(*c*), dependant on a constant *c*, such that :

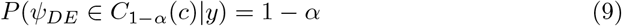

To build the confidence region, we also want to define it as to include regions of *ψ*_*DE*_ that have the highest density for the posterior distribution. *C*_1 *− α*_(*c*) is then defined as the Highest Density Region (HDR) [11]:

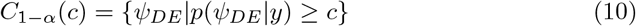

We define the null hypothesis *H*_0_ and alternative hypothesis *H*_1_ of our test, denoting 0 the null vector of dimension *N* :

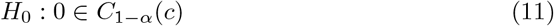

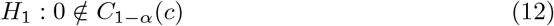

Using equations 9 and 10, the constant *c*^∗^ is defined as satisfying both conditions. In practice, both equations can be re-written such that:

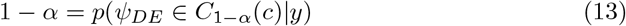

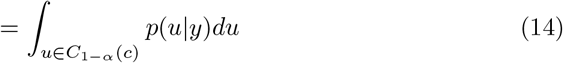

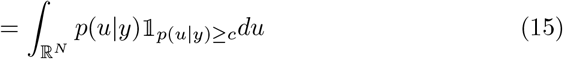

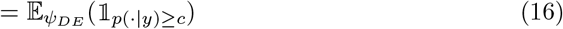

The last term of the equation can be approximated by a Monte Carlo procedure using the samples of *ψ*_*DE*_:

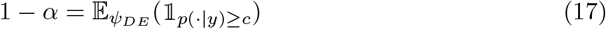

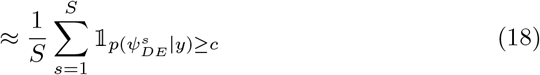

Finally, given that we can sample from *p*(·|*y*), *c*^∗^ can be approximated as the quantile of level *α* of the distribution of the 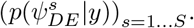.

To calculate *p*(·| *y*) from samples of *ψ*_*DE*_ from its marginal distribution, we use the assumption that all *N* patients are independent:

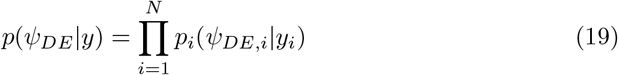

Each marginal density *p*_*i*_(·|*y*_*i*_) is a univariate density, and it can be estimated for example with the samples (*ψ*^*s*^ )_*s*=1…*S*_ using Kernel Density Estimation [26]. Finally, the test of the significance of the drug effect can assessed by testing if the vector 0 is inside the CR-HDR. We can reject *H*_0_ if:

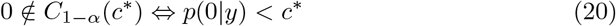

Finally, to correct the power of the test for the type I error, we choose the risk *α* such that the type I error would be 5%.

## C Pareto Depth Analysis method

For the construction of the Pareto Fronts, which is detailed in [7, 8], we consider the *M* individual features extracted from the data *B*_*i*_ (vector of dimension *M* ) with *P*_*train*_ the number of patients in the training set (patients simulated under *H*_0_) and *B*_*train*_ a *P*_*train*_ × *M* matrix containing the *M* -vector of features:

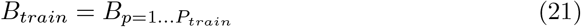

We then compute the set of dyads defined as the set of element-wise distances between each *B*_*train*_:

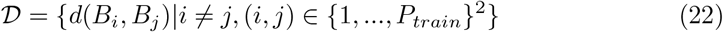

such that *d* is the element-wise distance:

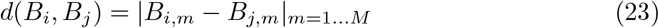

We define a domination relation ≻ such that *x* strictly dominates *y* (*x* ≻ *y*) if and only if:

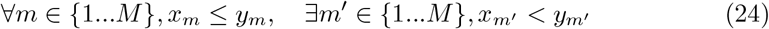

The first Pareto Front F_1_ is defined as the set of dyads in D which are not strictly dominated by any other dyads in *D*.

Iteratively, we build the *L*-th Pareto front as the sets of dyads in 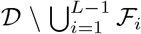 which are not strictly dominated by any dyad in 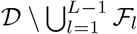.

In the end, each dyad is exactly in one Pareto front. Then we say that a dyad *D*_*o*_ is *below* a front ℱ_*l*_ if and only if:

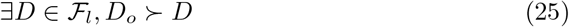

We finally define the *depth e*_*o*_ of *D*_*o*_ such that:

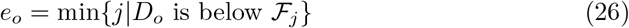

The full workflow is as follows:

With *B*_*train*_, calculate the set *D*, then construct the set of Pareto Fronts until each dyad is in a front.

Then, to test *N* observed patients, whose extracted features are 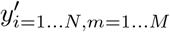 (a *N* ×*M* matrix), for each patient *i* ∈ {1…*N* } and each feature *m* ∈ {1…*M* }, we select the *k*_*i,m*_ nearest neighbours of 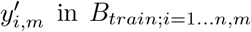 using the dissimilarity *d* (potentially with repetition) and evaluate the dyads. In the end, we have 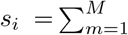 *k*_*i,m*_ new dyads.

*k*_*i,m*_ is selected independently for each feature and tested patient on the training set by the following procedure:

- initialize 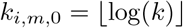
- increment *k*_*i,m*_ until the *k*_*i,m*_-NN graph of the training set (using *d* as the distance) is connected.
- evaluate the mean depth for the created dyads for each patient in *B*_*test*_, the *P*_*test*_ × *M* matrix containing the test patients simulated under *H*_0_:

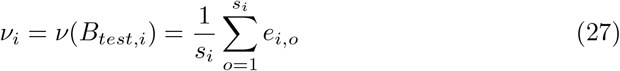

## D Modelling the SARA score in PROSPAX

This section describes model selection and fit in the PROSPAX dataset.

### Data

Table S-1 shows the characteristics of the population used in the analysis.

**Table S-1:** Characteristics of the PROSPAX patients for each genotype.

| Characteristics | ARSACS (n = 81) | SPG7 (n = 113) |
| --- | --- | --- |
| SARA score at inclusion, median (IQR) | 17.5 (13 - 22.5) | 11.5 (8 - 13.5) |
| Time Since disease Onset at inclusion, median (IQR), year | 30 (20 - 38) | 16 (10 - 23) |
| Age at inclusion, median (IQR), years | 37 (28 - 42) | 55 (47 - 61) |
| Female, n (%) | 42 (52) | 41 (36) |
| Age of Onset, median (IQR), years | 2 (1.5 - 7) | 38 (30 - 44) |
| Number of visits, n (%) |  |  |
| 1 | 12 (15) | 10 (9) |
| 2 | 21 (26) | 11 (10) |
| 3 | 48 (59) | 92 (81) |

Figure S-1 shows the SARA score versus the Time Since disease Onset (TSO) for both genotypes.

**Fig. S-1:**
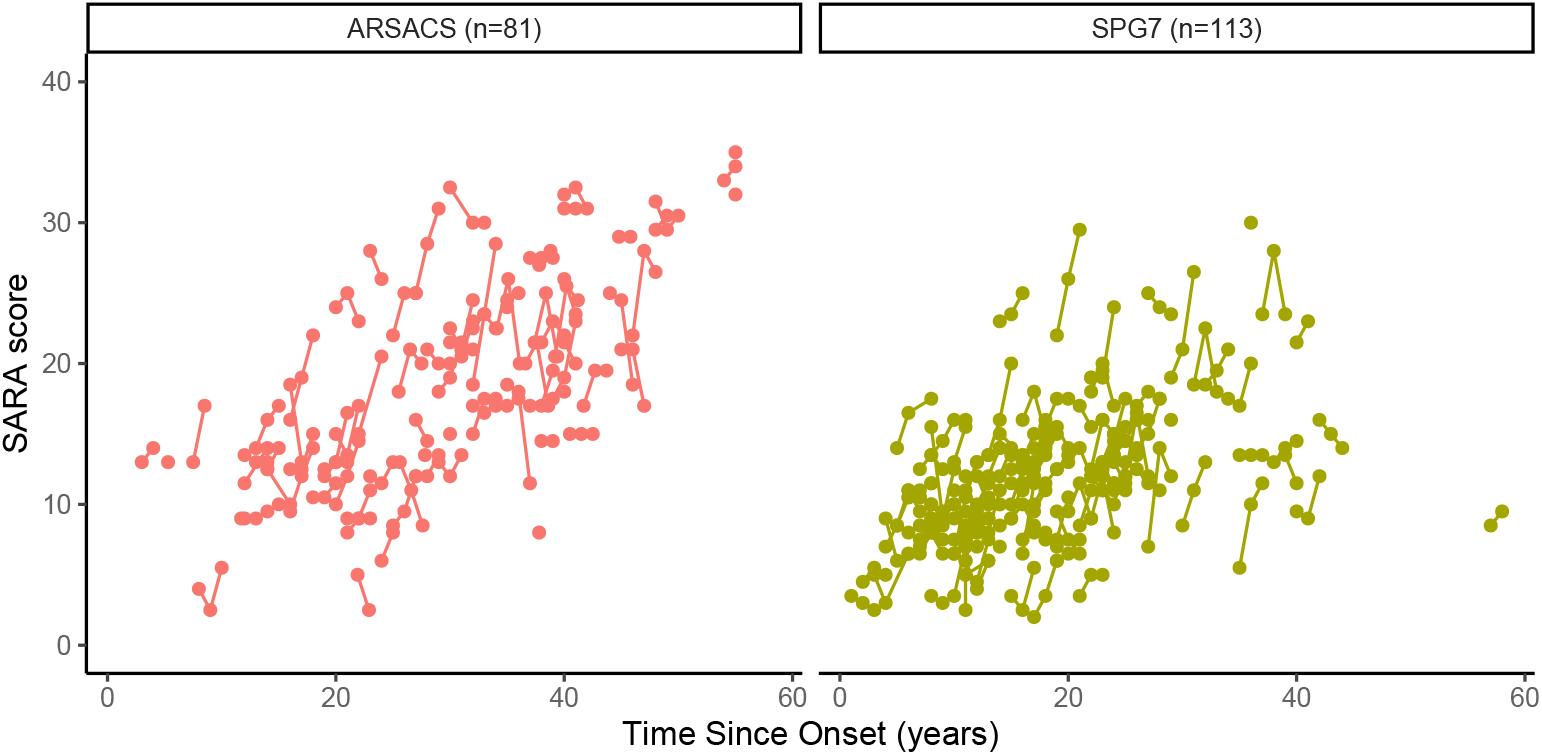
Plot of the SARA score as a function of TSO (Time Since disease Onset) for the ARSACS and SPG7 genotypes in the PROSPAX study. Solid lines represent the repeated measures for an individual.

### Methods

We analysed separately the two genotypes using the four-parameter logistic model in equation 1. The structure of random effects was selected according to a backward Likelihood Ratio Test (LRT) procedure (p-value=0.05). For the linear model, the covariate effects were pre-specified. The population parameters were estimated using the *saemix* package [13] in R 4.1.2 [14].

### Results

For both genotypes, variability on *γ* was removed from the model and for SPG7 we also removed the variability on *δ* and set this parameter to 0. Table 1 shows a good estimation of the population parameters, as the relative standard errors are below 50%. Diagnostic fits (Figures S-2a and S-2b) show adequate model fit.

**Fig. S-2:**
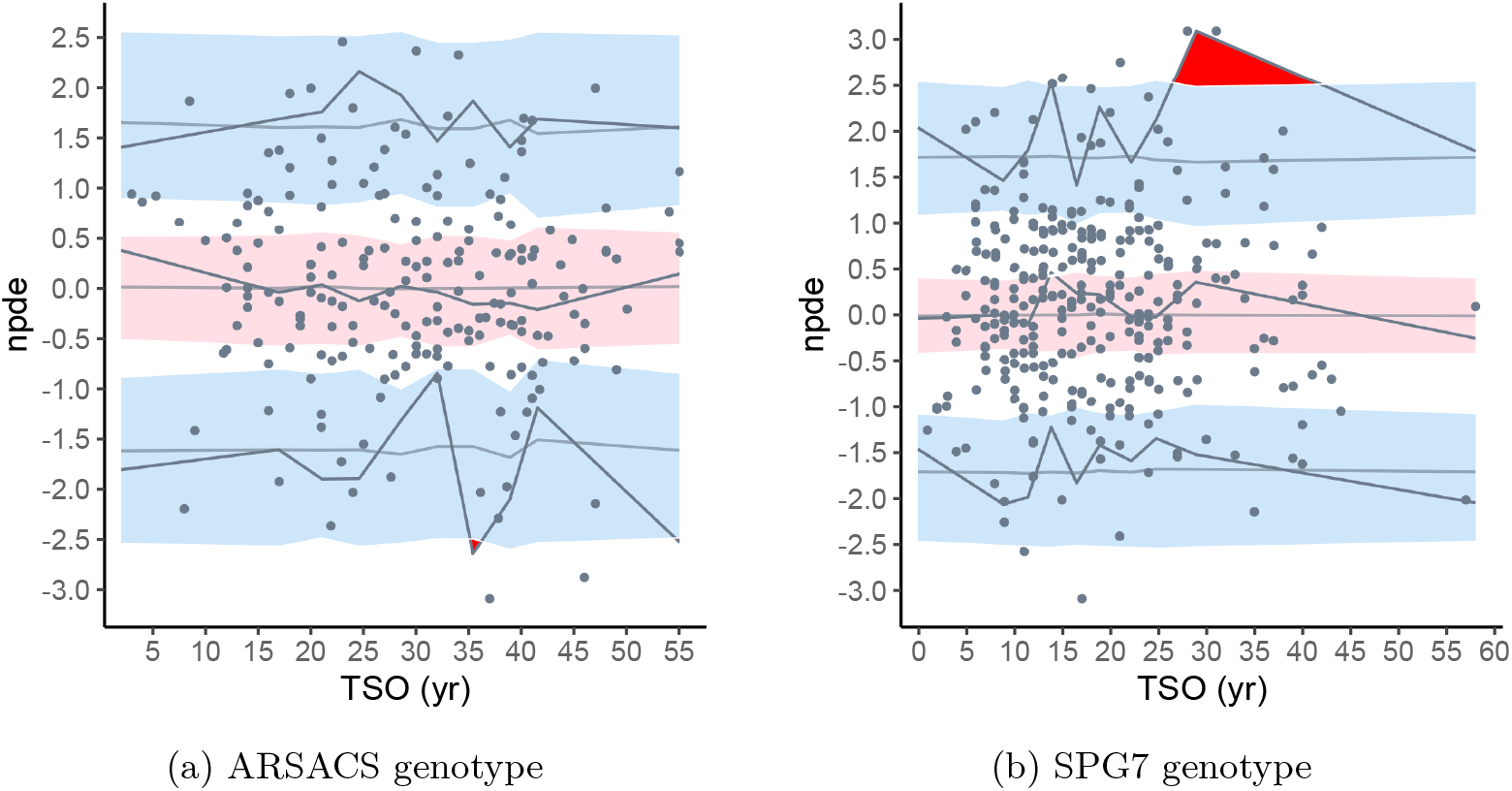
NPDE (Normalised Prediction Distribution Errors) versus Time Since disease Onset (TSO) of the logistic model fitted on each genotypes. The observed 5%, 50% and 95% percentiles are represented as solid black lines, the red band represents the simulated (1000 replicates) median npd with its 90% prediction interval, the blue line represents the simulated 5% and 95% percentiles (1000 replicates) with their 90% prediction intervals

## E CDDE method: Tables comparing the Wald and CR-HDR tests

Table S-2 shows the type I error and corrected power for the two tests performed with the CDDE method, for N-of-1 trials. Table S-3 shows the corresponding results for N-of-5 trials.

**Table S-2:**
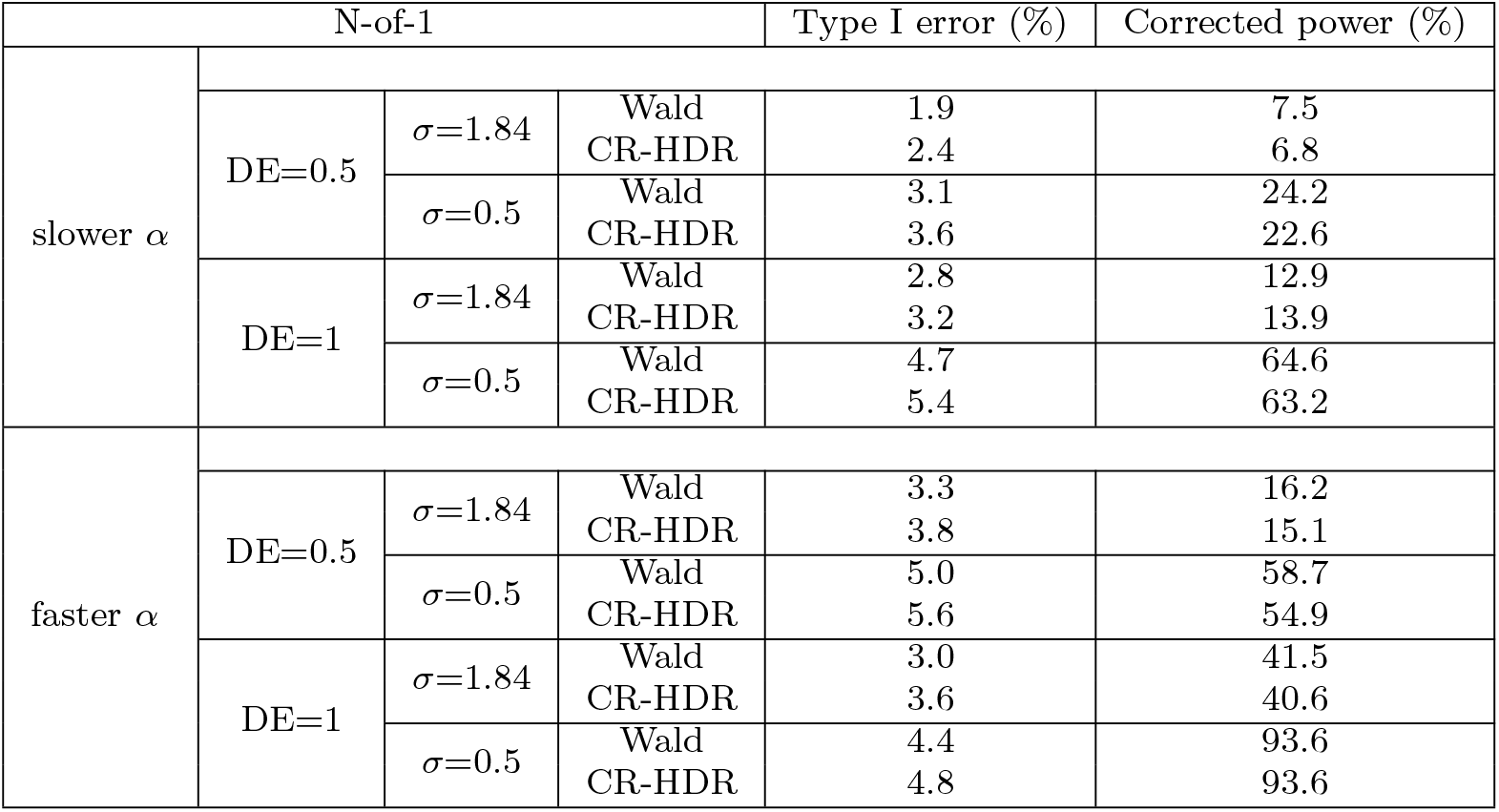
Table of the type I error and corrected power of the CDDE method, using the Wald test and the CR-HDR test for all N-of-1 scenarios, bold values correspond to inflated type I error.

**Table S-3:**
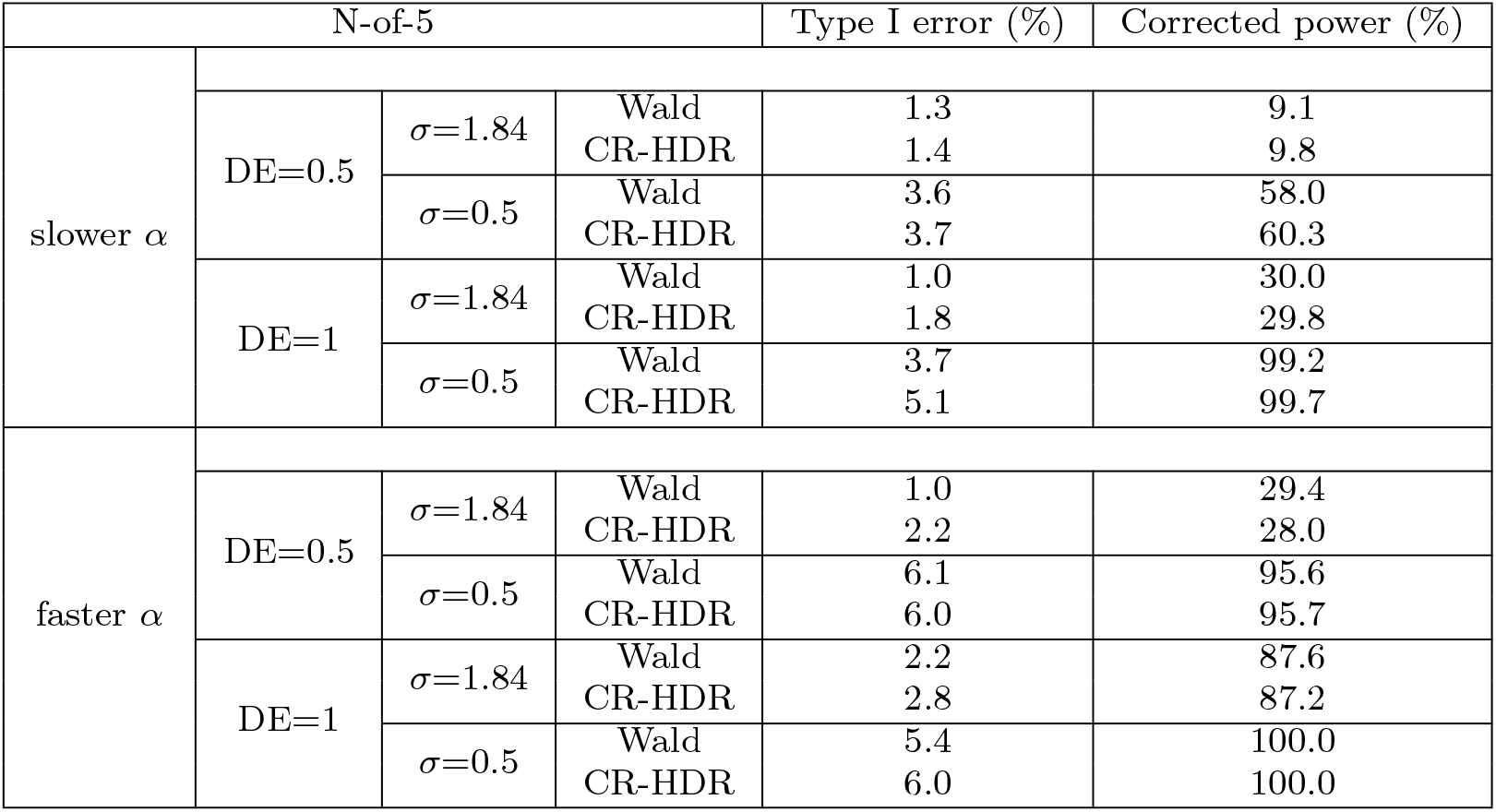
Table of the type I error and corrected power of the CDDE method, using the Wald test and the CR-HDR test for all N-of-5 scenarios, bold values correspond to inflated type I error.

**Table S-4:**
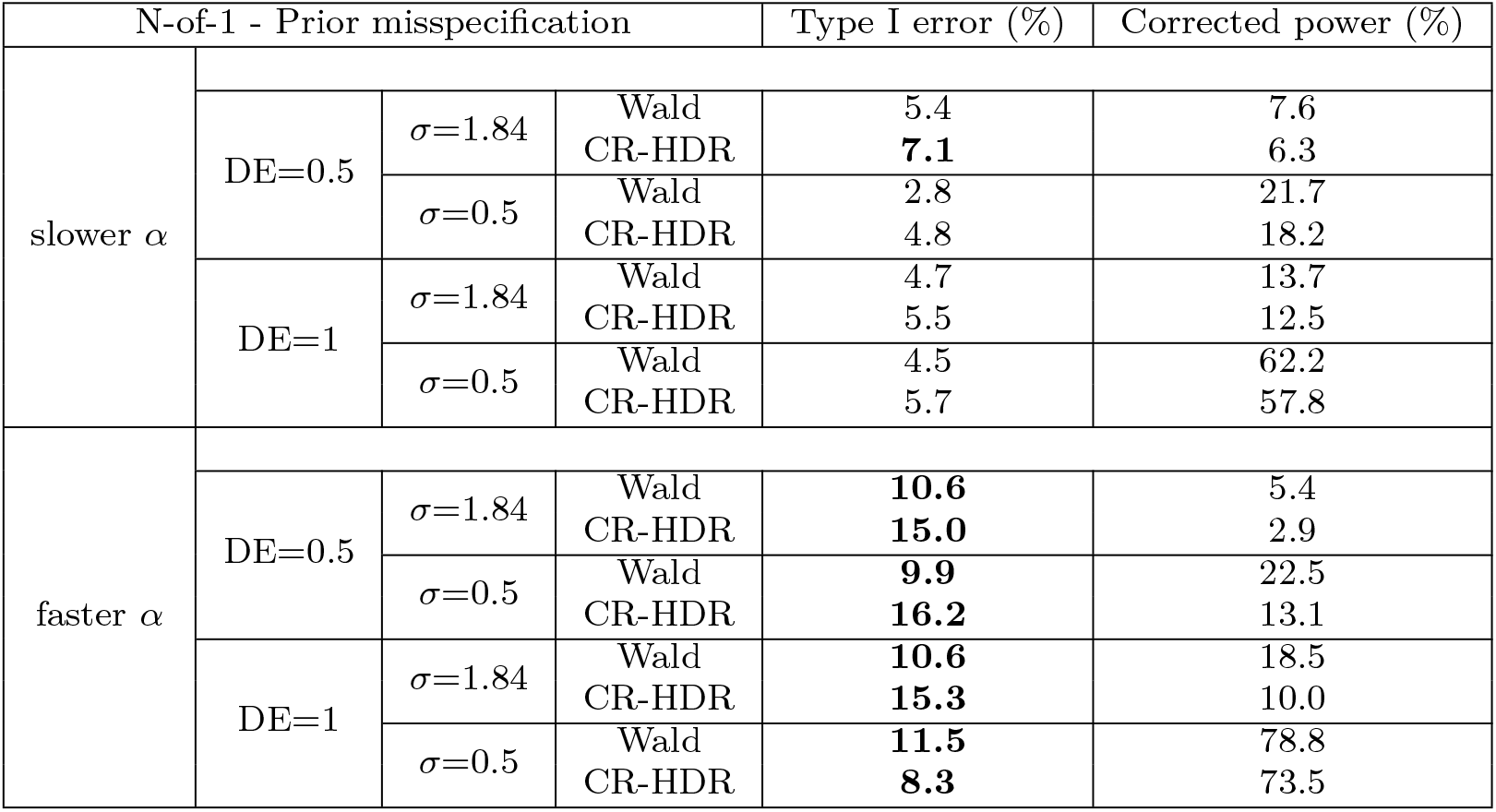
Table of the type I error and corrected power of the CDDE method of the prior misspecification scenarios, using the Wald test and the CR-HDR test, bold values correspond to inflated type I error.

**Table S-5:**
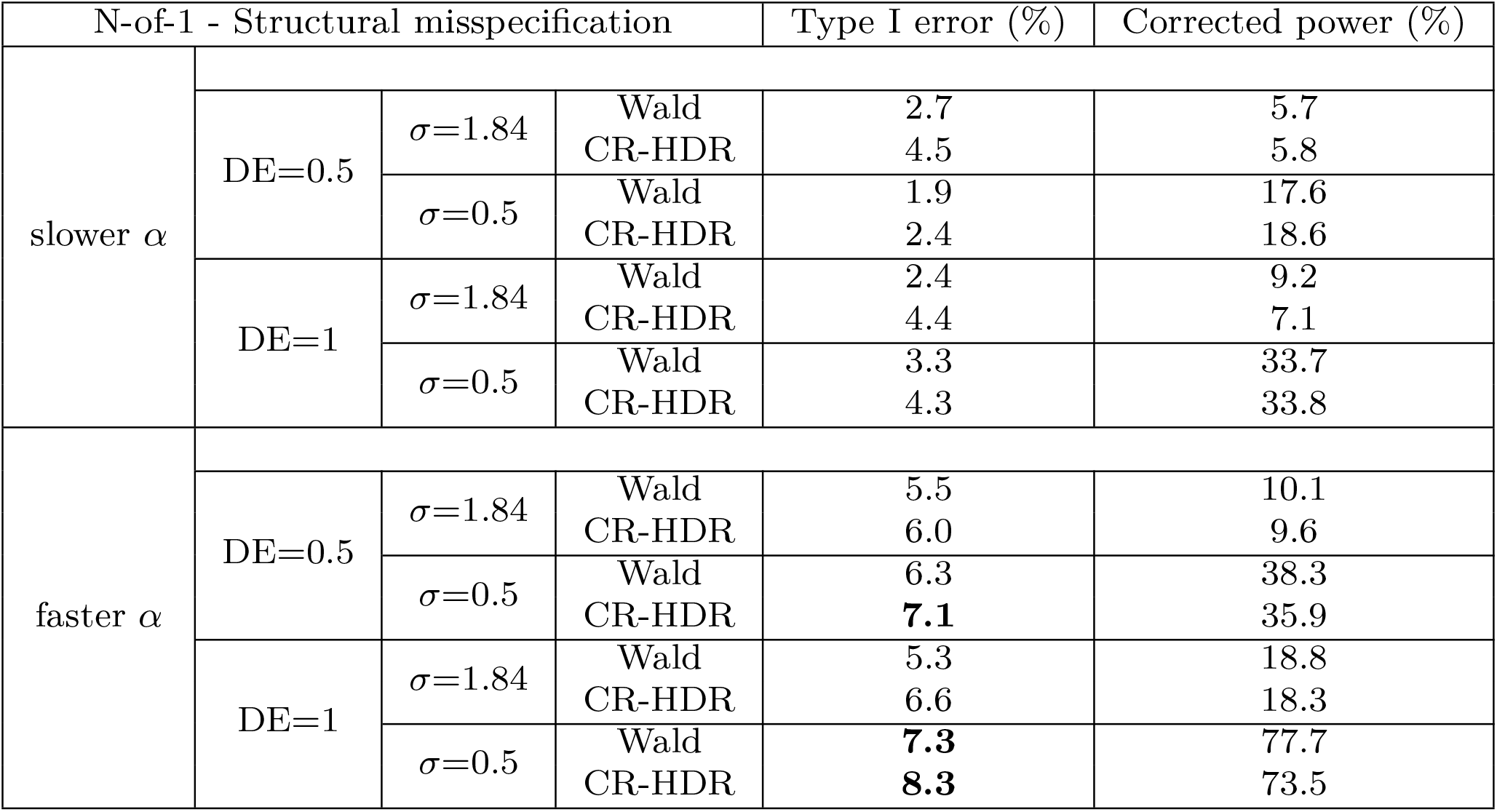
Table of the type I error and corrected power of the CDDE method of the structural misspecification scenarios, using the Wald test and the CR-HDR test, bold values correspond to inflated type I error.

